# Evaluating strategies to combat a major syphilis outbreak in Australia among Aboriginal and Torres Strait Islander peoples in remote and regional Australia through mathematical modelling

**DOI:** 10.1101/2021.03.07.21253089

**Authors:** Ben B. Hui, James S. Ward, Rebecca Guy, Matthew G. Law, Richard T. Gray, David G. Regan

## Abstract

**Background:** An ongoing infectious syphilis outbreak, first reported among Australian Aboriginal and Torres Strait Islander people in 2011, has resulted in >3000 notifications to the end of 2019 with multiple congenital syphilis cases and infant deaths. In 2017, the Australian Government introduced an enhanced test and treat response. We evaluate the impact of this response and the potential impact of further expansion of testing interventions.

**Methods:** We developed a mathematical model to capture the transmission of infectious syphilis among young heterosexual Indigenous Australians aged 15-29 years living in regional and remote areas. We used the model to assess the impact of existing and hypothetical outbreak responses on infectious syphilis prevalence.

**Findings:** The increase in testing coverage achieved through the enhanced response (18% coverage in 2011, to 39% in 2019) could lead to a stabilisation of the epidemic from 2021. To return to the pre-outbreak level (<0·2%) within five years, testing coverage needs to reach 60%. With the addition of a biannual community-wide screening program, using outreach to test 30% of youth in communities over 6 weeks,, a return to pre-outbreak levels can be achieved within 2 years. If testing coverage alone was scaled-up to 60% at the start of outbreak in 2011, syphilis prevalence would have returned to pre-outbreak levels by 2014.

**Interpretation:** Modelling suggests that to control the syphilis outbreak the response needs to be delivered with further potency. The reduction in prevalence could be hastened with community-wide screening at similar time periods across all communities along with increases in annual testing coverage.

**Funding:** The research was undertaken by the Kirby Institute, UNSW Sydney, for the Multi-jurisdictional Syphilis Outbreak Working Group (MJSO) with funding from the Australian Department of Health.

**Research in context:** *Evidence before this study:* We search PubMed with the terms ((“syphilis”[MeSH Terms] OR “syphilis”[All Fields]) AND (“disease outbreaks”[MeSH Terms] OR (“disease”[All Fields] AND “outbreaks”[All Fields]) OR “disease outbreaks”[All Fields] OR “outbreak”[All Fields])) AND “model “[All Fields]) on 12 March 2020 and identified 27 articles. Most articles focused on men who have sex with men and/or populations with HIV co-infection, neither of which is common in our target population (Indigenous Australians: predominantly aged 15-29; heterosexual and living in regional and remote areas). Of the remaining articles that consider syphilis control through screening interventions, the most relevant paper to our study is a modelling paper by Pourbohloul *et al*. in 2003, which demonstrated that community-wide treatment has no lasting effect on syphilis transmission.

*Added value of this study:* We developed a mathematical model to assess the impact of an enhanced response to a major syphilis outbreak in remote Aboriginal and Torres Strait Islander populations of Australia and whether it can be controlled by increasing testing coverage. The model captures sexual behaviour information and short-term population mobility patterns across regional and remote communities of Australia and was calibrated against the most recent infectious syphilis notification data to-date (up to 2019) and testing coverage data from the affected regions. Our findings provide an insight into the role of increasing testing coverage in controlling syphilis outbreaks among populations living in remote communities globally.

*Implications of all the available evidence:* Existing evidence suggests that enhanced surveillance, expanded clinical and laboratory services, enhanced health promotion, strengthened community involvement and a rapid outbreak response are core components to controlling syphilis outbreaks. Our study focuses on modelling the impact of expanded clinical services and ability to expand testing among the target population. Our study suggests that increasing testing coverage of the Aboriginal and Torres Strait Islander population aged 15-29 living in remote and regional communities to a level of 60%, would stabilise the epidemic and reduce overall prevalence to pre outbreak levels in around five years. Combining the 60% testing coverage with bi-annual community screening over a 6-week period involving outreach (minimum coverage of 30%) would reduce the time period to around 2 years. We believe these findings have implications for other Indigenous populations across the world who often live in remote regions with limited access to healthcare and are disproportionately affected by STIs.

## Introduction

While globally, the recent increases in syphilis prevalence have primarily occurred among men who have sex with men,^1,2^ an increase in infectious and congenital syphilis among the Indigenous heterosexual population in Australia and New Zealand has also been observed.^3-5^ This is of concern as many Indigenous groups face poor access to healthcare and are at risk of delayed treatment and the development of secondary diseases. In 2011, an outbreak of infectious syphilis among young Aboriginal and Torres Strait Islander people was detected in the north-west region of Queensland, Australia. The epidemic has since spread to other states and territories and has led into an endemic level of prevalence in some areas. As of September 2020, more than 3200 notifications had been reported, including 9 confirmed cases of congenital syphilis (corresponding to a rate almost 20 times higher than for the non-Indigenous population), and 3 deaths.^6,7^ Overall, 53% of cases have been diagnosed among women and 47% among men; 59% of all cases have been diagnosed in people aged 15-29 years.

Given the spread across jurisdictions in April 2015 a Multijurisdictional Syphilis Outbreak (MJSO) Working Group was formed to co-ordinate outbreak control efforts. The MJSO focussed on enhanced surveillance, strategies to increase testing of young people, including increasing testing coverage and frequency among pregnant women, timely treatment and contact management through primary health care clinics serving Aboriginal and Torres Strait Islander peoples.^8^ Concurrently a multi-pronged health promotion campaign was implemented to raise awareness about syphilis and to encourage young people to test. In December 2017, the Australian Health Protection Principal Committee (AHPPC) Governance Group developed a National strategic approach for an enhanced response to the disproportionately high rates of syphilis in Aboriginal and Torres Strait Islander people, which included expand testing coverage and the rollout of rapid point of care testing kits. Between 2014 and 2018 there was a notable increase in syphilis testing by as much as 1·8-fold in afflicted jurisdictions.^9^ However, during the same period, a rapid increase in infectious syphilis notifications was reported.^4^

We conducted mathematical modelling to assess the potential impact of syphilis testing on the epidemiology of the outbreak once it has become widespread and endemic. Specifically, we assessed the impact of the increase in testing coverage has achieved so far, the predicted impact on infectious syphilis prevalence should the testing coverage continue at the current level or increase further, and what testing coverage would be required to achieve a return to pre-outbreak syphilis infection rates. It has also been proposed that community-wide testing, where a large proportion of the community is tested and treated over a short period achieved through resource intensive and well coordination outreach strategies, may be an effective intervention for syphilis control. While community-wide testing in isolation is not likely to be effective in the long term,^10^ it has the benefit of reducing the prevalence of infection within a short period thereby increasing the benefits of ongoing clinic-based testing and treatment, and is thought to be feasible for implementation in some remote communities. We therefore investigated the impact of community-wide testing as a stand-alone strategy in remote communities and the potential impact if implemented concurrently with increased testing coverage, via routine testing youth at primary care services. Finally, we investigated the impact of an early scale-up of syphilis testing once the outbreak was detected to see if its spread could have been prevented or substantially mitigated.

## Methods

### Governance and engagement

Since 2015 a multijurisdictional syphilis outbreak committee has provided governance oversight to the outbreak response and in 2017 the Federal Government introduced an enhanced test and treat strategy. The MJSO Working Group consisting representatives from affected jurisdictions, sexual health physicians, experts in Aboriginal and Torres Strait Islander sexual health and the Australian Government Department of Health engaged us to assess the trajectory of the syphilis outbreak, assess the potential impact of the response, and determine the testing coverage required to return syphilis incidence and prevalence to pre-outbreak levels. The Working Group reviewed and provided oversight of this work, provided syphilis notifications and testing data, and informed the develop of the scenarios we investigated.

### Model overview

We extended a previously developed individual-based simulation model of sexually transmitted infections (STIs) in heterosexual Indigenous Australians to capture the syphilis outbreak across the regions affected by the outbreak.^11^ The model tracks the sexual transmission of syphilis among a population of 10,550 males and females, aged 15-34 years, representing one regional community setting (of 5,550 people) and 10 smaller remote communities (of 500 people each), respectively. The sex and age distribution of the modelled populations is based on Indigenous Australians’ demographic data obtained from the Australian Bureau of Statistics.^12^ We used this population structure to reflect available demographic data and to represent the overall Indigenous population living in typical syphilis outbreak areas. A summary of the modelling methodology follows, with the accompanying Supplementary Material providing full implementation details with model code available online.^13^

Individuals in the model have a designated “home” location but can move between a regional community and remote communities according to the estimated proportion of individuals away from home reported by Biddle and Prout.^14^

Individuals can form sexual partnerships while they are at the same physical location. They can also be sexually active while travelling or away from home, forming casual partnerships, even if partnered. The frequency of partner change and duration of partnerships is determined by the partner acquisition rate calculated based on the number of partners in the last 12 months as reported from a study on the sexual behaviour of remote Indigenous Australians.^15^

The natural history of syphilis is captured in the model in an analogous fashion to that adopted in a previously published modelling study of syphilis transmission in an MSM population and as adopted by Gray *et al*., ^16-22^ and is illustrated schematically in Figure 1: Stages of disease progression for syphilis as captured in the model. Dashed arrows represent the additional pathways for infected individuals who have received treatment.. Infected individuals are infectious if they are in the exposed, primary, secondary, early latent, or recurrent stages of syphilis. If left untreated, individuals will eventually develop tertiary syphilis which will only resolve when they receive treatment. The model assumes an infected person who receives treatment at or after reaching the latent stage will be immune from re-infection for 5 years on average, whereas no immunity is gained if an infected person is treated prior to the latent stage. The duration of infection and the transmission probability per sex act varies depending on the infection stage and the assumed values for these are listed in the Supplementary Material.

**Figure 1:**
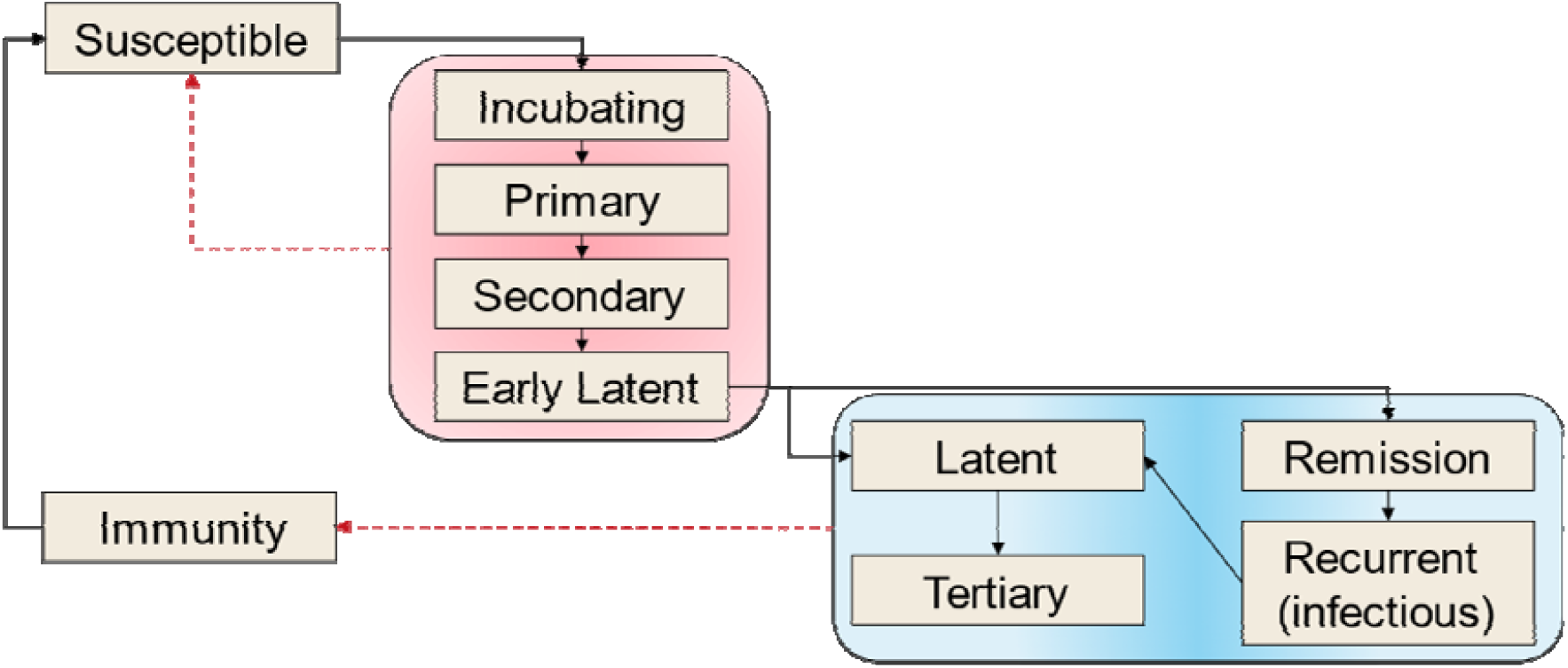
Stages of disease progression for syphilis as captured in the model. Dashed arrows represent the additional pathways for infected individuals who have received treatment.

### Syphilis testing

We assume ongoing syphilis testing of individuals is carried out in the model population. We used the overall number of tests and the estimated resident population size in the outbreak affected regions (QLD, NT, SA, and WA) between 2013-2019, provided to us by the MJSO, to estimate a crude population testing coverage.^9^ We then adjusted this coverage to reflect a likely overestimation of the numerator —due to including tests conducted in visitors from outside the outbreak affected areas, and multiple tests in individuals (such as from pregnant women)—and uncertainty in the population size (denominator). Based on the available data (described in the Supplementary Material) we multiplied the crude testing coverage by 0·75 to produce a change in testing coverage from 24% in 2013 to 39% in 2019. We assumed a varying testing coverage based on sex (with coverage lower in males than in females), region (with coverage in remote areas higher than in regional centres) and age (15-19, 20-24, 25-29, 30-34 years old, with coverage decreasing as age increases). For the period prior to 2013, we assumed a constant population testing coverage of 18% as reported in a previous study.^23^

We assumed that syphilis is detected through testing using assays with a sensitivity of 98%.^24^ Following diagnosis, all people are treated within 7 days in regional areas and 85% of people are treated within 4 months in remote areas based on the gonorrhoea and chlamydia treatment rates, and 15% are untreated.^25^ In the model we assumed there is no treatment failure and that people become susceptible again to re-infection following treatment.

We used the model to evaluate a range of scenarios involving increased annual testing (testing at least once during the year) coverage and community-wide testing (testing a large proportion of the community in a short period of time which for this study we specified as 30% of people aged 15-34 in each community to be tested for syphilis within 6 weeks). The scenarios were developed in consultation with the MJSO Working Group and jurisdictional stakeholders to reflect what we consider to be a feasible expansion of current intervention efforts to combat the syphilis outbreak in the affected areas. A brief description of each scenario is provided in Table 1: Syphilis testing scenarios considered in this study.. The interventions investigated with these scenarios are focused on increases in testing and treatment in response to syphilis becoming widespread and endemic. We do not consider other interventions such as contact tracing which can have a substantial impact when outbreaks are small and clustered.

**Table 1:** Syphilis testing scenarios considered in this study.

| Scenario | Description |
| --- | --- |
| 0 | Counterfactual scenario where there is no increase in testing coverage since the start of syphilis outbreak in 2013 and testing coverage remains at 2013 levels. |
| 1 | Annual testing coverage (39%) remains at the levels reported for the end of 2019 across all populations and regions. (baseline) |
| 2 | Annual testing coverage is increased from baseline level to 60% across all populations and regions over a period of 2 years. |
| 3 | Annual testing coverage is increased from baseline level to 60% across all populations and regions over a period of 5 years. |
| 4 | Annual baseline testing coverage is maintained and in addition, at the end of first two years, community-wide testing is implemented in remote locations whereby 30% of the community is tested over a 6-week period. Community-wide testing starts and finishes in all locations at the same time. |
| 5 | Annual testing coverage is increased from baseline level to 60% across all populations and regions over a period of 2 years. In addition, at the end of first two years, community-wide testing is implemented in remote locations whereby 30% of the community is tested over a 6-week period. Community-wide testing starts and finishes in all locations at the same time. |

In Scenario 5, community-wide testing starts and finishes at the same time in all remote locations, which could be difficult to operationalise. Therefore, we also investigated the case where community-wide testing screening starts and finishes at different times for different remote locations. Here, we assume, on top of increasing annual testing coverage of population (to 60% within 2 years), all remote locations can start community-wide testing (30% of the population over a six-week period) within the first year, but not all locations can start and finish at the same time. Under this assumption, we investigate the cases where: a) one, two and five remote locations start community-wide testing at the same time, with starting time for each round of community-wide testing spread out evenly throughout the year, and b) five remote locations can start community-wide testing at the same time (hence 2 rounds in total), with various time delay (from 30 days to 330 days) between two rounds of testing. We compare the model outcomes with the scenario where all community-wide testing can start at the same time (i.e., Scenario 5).

We also use the model to assess the potential impact of scaling up testing from 2011 (when the outbreak was first detected) to the level specified in Scenario 2 rather than from the end of 2019. This analysis is conducted to assess if earlier scale-up of wide-spread testing could have prevented the outbreak.

### Model outcomes

For each scenario results are collated for 100 simulation runs (or model realisations). Initially, to establish a baseline, the model was run 5000 times, and the 100 runs with the closest match to the 6-monthly infectious syphilis notification rate reported by the MJSO over 2013-2019 were selected (those with the smallest summed squared differences between model output and data).^7^ A more comprehensive description of this process is provided in the Supplementary Material.

The primary outcome of the model that we report is the prevalence of infectious syphilis in the population up to year 2030, the date when infectious syphilis prevalence returns to the pre-outbreak level (0·2%), and the date when syphilis prevalence reaches zero (which we refer to as elimination). Results are described by the median of the monthly estimates from the 100 runs with the uncertainty specified by the Inter-Quartile Range (IQR).

## Results

Figure 2 shows diagnoses per 100,000 population for the counterfactual and baseline scenarios (Scenario 0 and Scenario 1, respectively) over the period 2010-2025. These results suggest that if baseline testing rates can be maintained in outbreak affected areas, it has the potential to stabilise the outbreak post 2021. Assuming the annual testing coverage reached by end-2019 remains constant at 39% (Scenario 1). In Figure 2, we compare the diagnosis rates in the baseline and counterfactual scenarios to show what might have been seen by the surveillance system. For the baseline scenario the increase in testing results in a higher diagnosis rate but it then stabilises as treatment results in a lower prevalence the model predicts that the diagnosis rate (average calculated for the preceding 6 months) will remain relatively stable from the end of 2019 (341·2 per 100,000; IQR: 246·5 - 398·1) to the start of 2025 (360·2 per 100,000 IQR: 189·6 – 597·2). In contrast, if the annual syphilis testing coverage remained at the lower 2013 level of 24% (Scenario 0), then the estimated diagnoses rate for the previous 6 months will initially be lower than the baseline scenario (as the lower testing rate finds fewer infections) at 265 (IQR: 190 – 408) per 100,000 people at the end of 2019, but then increase rapidly to 588 (IQR: 398 – 730) per 100,000 people by 2025.

**Figure 2:**
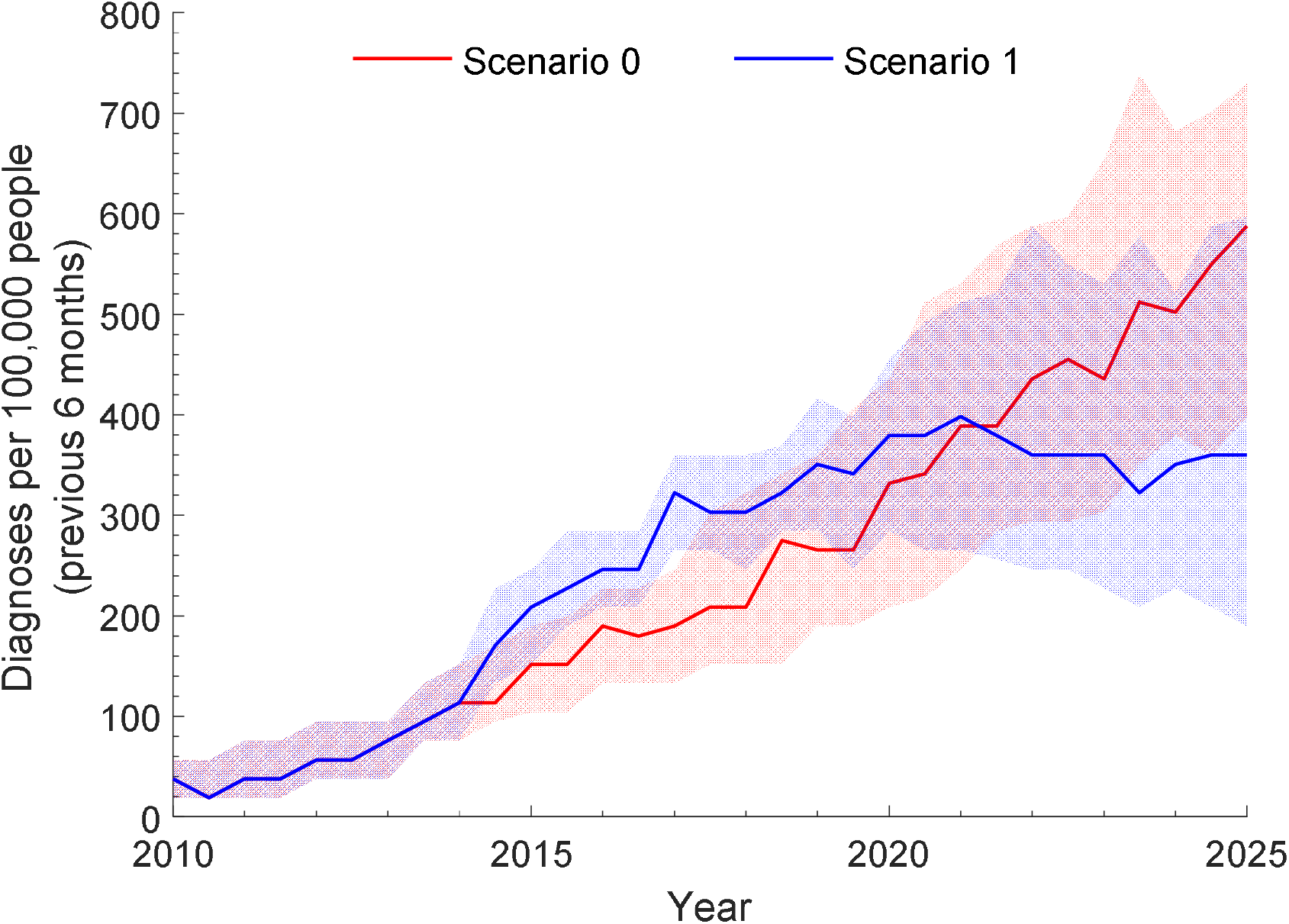
Diagnoses per 100,000 people from 2010 to 2025 for the counterfactual scenario (Scenario 0 in red; where testing rates remain at 2013 levels) and the baseline scenario (Scenario 1 in blue). The solid lines and shading are the median and interquartile range from the 100 selected model runs, respectively.

Under Scenario 0, infectious syphilis prevalence is predicted to increase from 0·38% (IQR: 0·27 – 0·51%) at the end of 2011 to 3·29% (IQR: 2·76 – 3·93%) by the end of 2025, while under Scenario 1 prevalence is only predicted to increase to 0·76% (IQR: 0·36 – 1·20%) over the same period.

Under Scenarios 2 and 3, we investigate the impact of increasing the annual testing coverage on infectious syphilis prevalence. From Figure 3 increases in annual syphilis testing coverage are predicted to substantially reduce infectious syphilis prevalence. If annual testing coverage increases from 39% to 60% by 2022 (Scenario 2), then infectious syphilis prevalence is predicted to decline to 0·14% (IQR: 0·03–0·25%) by 2026. If this increase in annual testing coverage is not achieved until 2025 (Scenario 3), the infectious syphilis prevalence would decline to 0·24% (IQR: 0·11–0·41%) by 2026. Increasing annual testing coverage to 60% is predicted to increase the probability of eliminating syphilis by 2044 from 31% (Scenario 1; baseline) to more than 90% under Scenarios 2 and 3, and to reduce the time required for elimination from 14 years to 8 years or 11 years, if the increase is achieved within 2 years (Scenario 2) or within 5 years (Scenario 3), respectively (Table 2). Under Scenarios 4 and 5 we investigate the impact of two annual rounds of community-wide testing in remote locations on infectious syphilis prevalence, either in isolation (Scenario 4) or in addition to increasing ongoing annual testing coverage (Scenario 5). Figure 3 shows community-wide syphilis testing as implemented in the model has only a small impact on syphilis prevalence when implemented in isolation. However, a much more dramatic reduction in infectious syphilis prevalence is predicted if community-wide testing is combined with increasing ongoing annual testing. In this case, prevalence declines to <0·1% by 2025 (Figure 3).

**Table 2:**
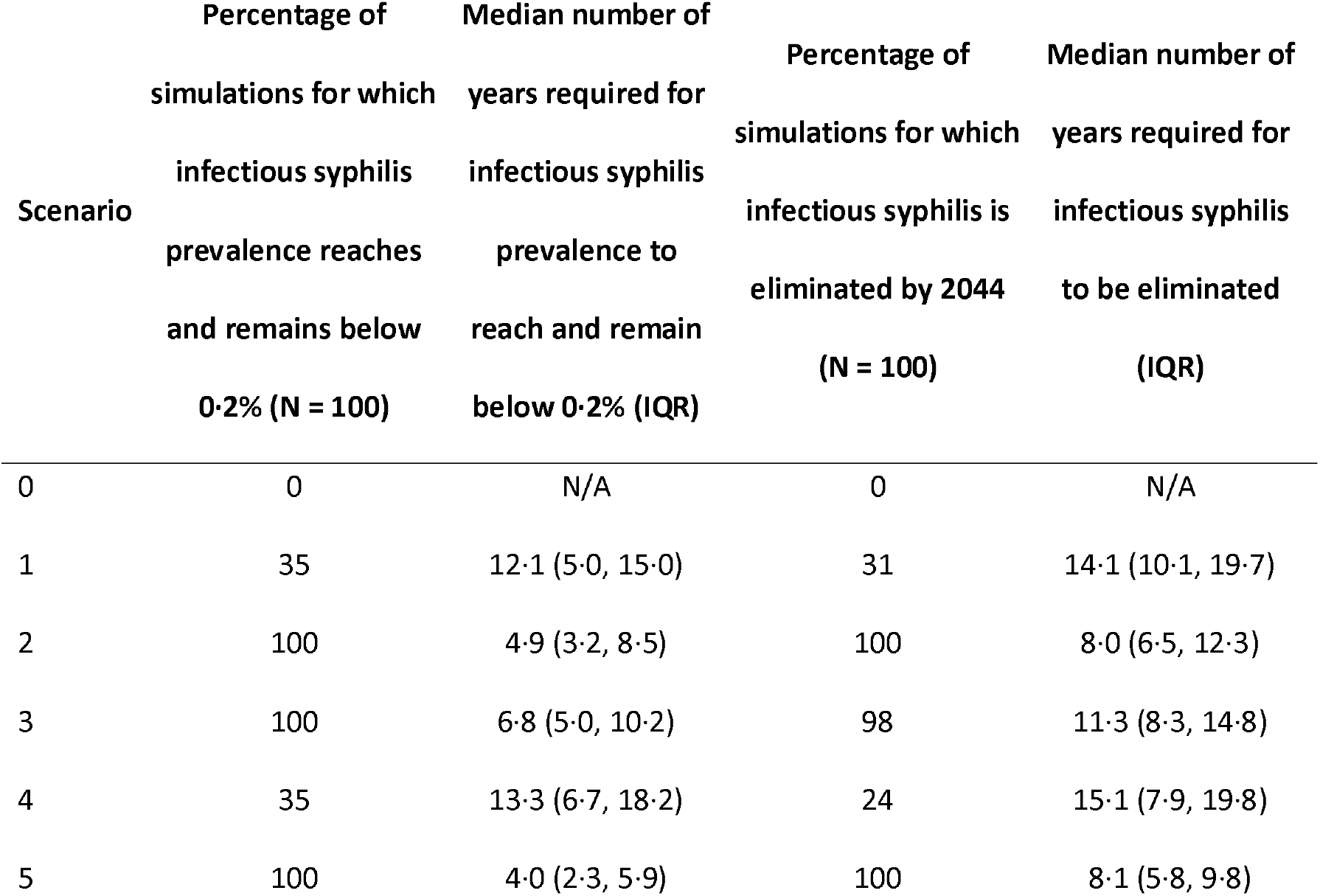
Percentage of simulations and time required for the infectious syphilis prevalence to reach and remain below 0·2% (columns 1 and 2, respectively), and infectious syphilis to be eliminated (columns 3 and 4, respectively), under each of the scenarios evaluated.

**Figure 3:**
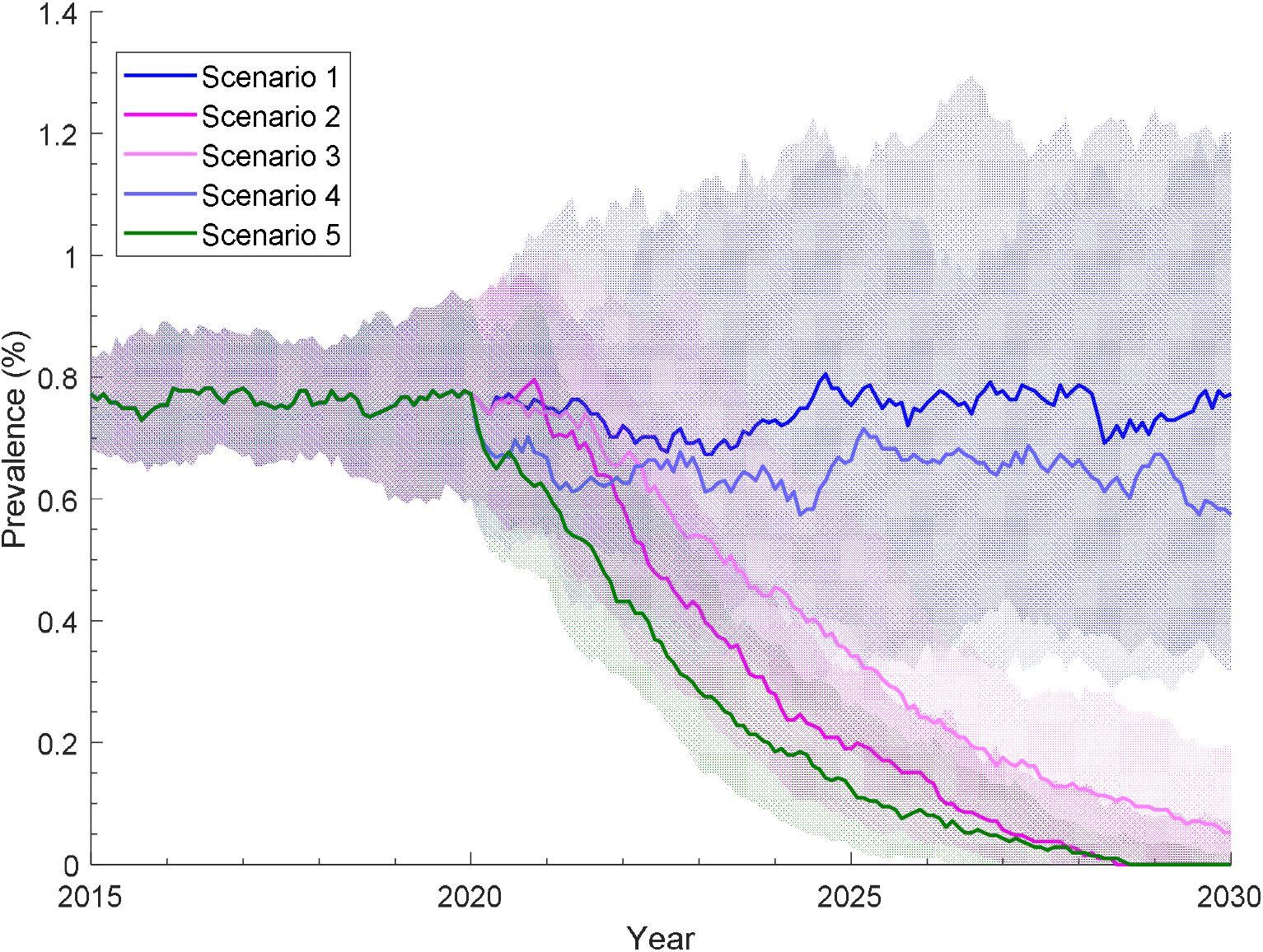
Infectious syphilis prevalence trajectories for Scenarios 1 −5 over the period 2015-2030. The solid lines and shading are the median and interquartile range, respectively, from 100 model runs that provide the closest match (in terms of smallest summed square differences) with the notification rate from 2013 to the end of 2019.

Scenario 2 above assumes increase of annual testing coverage starts from the end of 2019. In the Supplementary Material (Figure A7 and Table A9) we examine the benefits if such increase occurred at an earlier stage of the outbreak. If the testing coverage increases to 60% at the end of 2013 instead of 2019, syphilis prevalence would have been likely to return to below 0·2% by 2017, and by 2014 if the increase is introduced at the end of 2011. The peak prevalence and duration of the outbreak are reduced in both scenarios, with an additional 591 (if start at the end of 2013) and 702 (if start at the end of 2011) infections averted when compare to Scenario 2.

Table 3 and 4 summarise the time required for infectious syphilis prevalence to return to pre-outbreak level (defined as prevalence staying below 0·2%) and for elimination under increasing annual testing coverage and multiple rounds of community-wide testing. Table 3 summarises the results when one, two and five remote locations can start community-wide testing at the same time, with the starting time for each round of community-wide testing spread out evenly throughout the year. Table 4 summarises the results when five remote locations can start community-wide testing at the same time with time delay ranging from 30 days to 330 days.

**Table 3:** Time required for infectious syphilis prevalence to reach and remain below 0·2% (columns 4) and infectious syphilis to be eliminated (column 5) under multiple rounds of community-wide testing on top of increasing annual testing coverage (to 60% within 2 years, i.e. Scenario 4). Each round of testing consists of testing 30% of population at one or multiple remote locations over a period of 6 weeks, repeating annually for two years. Community-wide testing is spread out evenly such that by the end of first year all 10 remote communities will have experienced community-wide testing.

| **Number of rounds of population-wide testing** | **Number of remote locations included per round of community-wide testing** | **Delay between rounds of community-wide testing (days)** | **Median number of years required for infectious syphilis prevalence to reach and remain below 0.2% (IQR)** | **Median number of years required for infectious syphilis to be eliminated (IQR)** |
| --- | --- | --- | --- | --- |
| 1 | 10 | N/A | 4.0 (2.3, 5.9) | 8.1 (5.8, 9.8) |
| 2 | 5 | 180 | 5.2 (2.6, 7.1) | 9.5 (7.1, 11.9) |
| 5 | 2 | 72 | 4.5 (2.7, 6.8) | 8.5 (6.5, 11.7) |
| 10 | 1 | 36 | 4.4 (3.0, 6.7) | 8.3 (6.3, 10.8) |

**Table 4:** Time required for infectious syphilis prevalence to reach and remain below 0·2% (columns 2) and infectious syphilis to be eliminated (column 3) under two rounds of community-wide testing on top of increasing annual testing coverage (to 60% within 2 years, i.e., Scenario 4), with delay between rounds from 30 days to 330 days. Each round of testing consists of testing 30% of population at five remote locations over a period of 6 weeks, repeating annually for two years

| **Delay between the first and second rounds of community-wide testing (days)** | **Median number of years required for infectious syphilis prevalence to reach and remain below 0.2% (IQR)** | **Median number of years required for infectious syphilis to be eliminated (IQR)** |
| --- | --- | --- |
| 0 | 4.0 (2.3, 5.9) | 8.1 (5.8, 9.8) |
| 30 | 4.7 (3.0, 6.6) | 8.6 (6.8, 12.2) |
| 60 | 4.5 (2.7, 7.5) | 8.7 (6.8, 11.1) |
| 90 | 4.9 (2.5, 7.0) | 8.8 (6.4, 11.8) |
| 120 | 4.6 (2.7, 6.6) | 8.4 (5.6, 11.7) |
| 150 | 4.9 (3.0, 7.4) | 9.0 (6.2, 12.1) |
| 180 | 5.2 (2.6, 7.1) | 9.5 (7.1, 11.9) |
| 210 | 5.4 (3.1, 8.4) | 10.2 (7.1, 13.8) |
| 240 | 4.6 (2.9, 6.8) | 8.4 (6.3, 12.3) |
| 270 | 4.2 (3.2, 7.1) | 8.4 (6.8, 11.5) |
| 300 | 4.4 (3.0, 6.8) | 8.0 (6.0, 11.8) |
| 330 | 5.0 (3.0, 7.4) | 8.9 (6.8, 12.3) |

## Discussion

The findings from our modelling analysis suggest that the increase in testing that has been achieved since the beginning of the outbreak has led to a stabilisation of the epidemic and averted a substantially worse one (Scenario 0). At the current level of testing, our model predicts a 30% probability of eliminating infectious syphilis in the affected regions by 2044 and that elimination is almost certain if annual syphilis testing coverage is increased to 60%. If two annual rounds of community-wide testing in remote locations are conducted, in combination with an increase in annual testing coverage, the time required for infectious syphilis prevalence to return to the pre-outbreak can be brought forward by approximately one year. However, community-wide testing on its own does not reduce the time required for syphilis elimination and it has no incremental benefit in terms of reducing infectious syphilis prevalence beyond 2027, as the spread of infectious syphilis from regional centre to remote locations is not affected by community-wide testing.

In a previous modelling study by Oxman *et al*., it is suggested that a small group (as little as 0·1% of the total population) of individuals with high numbers of sexual partners (300-400 per year, or >8 per week) can trigger a syphilis epidemic.^26^ In comparison, among our selected simulations (i.e., simulations for which the notification rate matches the reported infectious notification rate from the start of the outbreak), the maximum number of partners in the last year for any individual is 6. Our findings suggest that syphilis epidemics can be sustained with much lower partner numbers than those predicted by Oxman *et al*. to be necessary, and the occurrence could depend on a range of stochastic (e.g., contact network and behaviour of the infected individual) and non-stochastic (e.g., population demographic and baseline testing coverage) factors ^28-30^.

While increasing annual testing coverage is likely to remain one of the key testing strategies for reducing transmission and lowering prevalence, community-wide testing in short time periods has the potential to lower prevalence faster than without. Under the assumption that community-wide testing is only feasible for smaller remote locations (i.e., not for larger regional centres), our findings suggest that, ideally, community-wide testing should be carried out at all locations at the same time if reducing prevalence quickly is the primary objective. Similar to the findings in Pourbohloul C *et al*. however, we also find that without concomitant increase in annual testing coverage, infectious syphilis prevalence rebounds quickly after community-wide testing and treatment with the time required for prevalence to return to the pre-outbreak level essentially the same as the time required if no community-wide testing is implemented.^10^

Given the logistical and economic challenges associated with the implementation of community-wide testing over vast geographical distances, we also investigate the effect of asynchronous community-wide testing across remote locations. Our result suggests there is no simple relationship between the delay and the time needed for infectious syphilis prevalence to return to pre-outbreak level. Instead, it depends on other complexities such as population mobility, the coverage of the community-wide testing, the duration of community-wide testing, and the likelihood of testing period overlapping. As a crude estimate, our result suggests that if the delay between the starts of community-wide testing is inevitable, then shorter the delay is usually better, with 50% of results led to a quicker return to pre-outbreak prevalence (when compare to equivalent scenario but without community-wide testing) if delay is less than 360 days, as oppose to 33% if delay is more than 360 days.

Unsurprisingly, if the scaling up of syphilis testing coverage across the population happened earlier during the outbreak, then our model predicts a much smaller outbreak with elimination to occur prior to 2019 (see Supplementary Material). It should be noted, however, that our model and scenarios were designed to assess the impact on a syphilis outbreak once it becomes established and has spread. It is likely the outbreak started within a small undetected cluster and then spread to and expanded within multiple regions. If this cluster was detected earlier with a faster initial response and the robust implementation of other public health interventions not investigated here, such as contact tracing, then the outbreak may not have become endemic across wide parts of Australia even if overall testing coverage remained low. This highlights the importance of a rapid and committed response to emerging outbreaks of syphilis globally.

Several limitations of our modelling approach must be considered in the interpretation of our findings. First, the model is calibrated in terms of notification and testing data from syphilis affected regions, with the affected regions included changing dynamically as the outbreak progressed. We also estimated the annual testing coverage after 2013 using the number of tests carried out and the estimated population size, while testing coverage prior to 2013 was assumed to be the same as the annual testing coverage for other STIs from a previous study.^23^ This means the testing coverage used in the study might not be a true representation of the actual syphilis testing coverage within a specific region through time. We assumed the number of syphilis tests reported overestimated the actual testing coverage due to a number of contributing factors (see Supplementary Material for details), and the testing coverage was adjusted during model calibration and in the baseline model. The sensitivity of this adjustment was assessed with additional results for various levels of adjustment provided in the Supplementary Material. Second, upon positive diagnosis, we assumed individuals are treated for syphilis within the same timeframe as reported for a positive diagnosis in gonorrhoea and chlamydia. It is possible treatment may have been initiated sooner in the outbreak affected areas due to the enhanced response to the syphilis epidemic. Third, this study only considered increased testing coverage as a response to the infectious syphilis outbreak and did not consider other effects of the enhanced response, such as changes in sexual behaviour that reduce the risk of acquisition due to the parallel rollout of communication and education materials across the outbreak affected regions, though recent data suggests an increase in partner exchange rate among high activity group.^31^ Fourth, we have previously shown that population movement between remote locations is an important factor in sustaining STI transmission^11^, but there are scant data to inform this and hence remains a source of uncertainty. Finally, the model was not designed to investigate or identify the cause of the outbreak, with simulations only to be included in this analysis if an outbreak has occurred and has led to notification rate over the 2013-2019 period. This selection process means the syphilis persistence shown in our baseline scenario (Scenario 1) does not fully show the stochasticity outputted by the model. As Figure A1 in the Supplementary Material shows, without the simulation selection process, syphilis does not persist in the majority of simulations. On a related note, at the time of writing, the overall notification rate across the entire outbreak region shows early signs of levelling off, and hence the simulation selection process is biased towards simulations where a further increase in notifications is unlikely. Therefore, our findings might not be applicable to all affected regions, as such a levelling off of the outbreak overall could mask a rising numbers of cases and emerging outbreaks in specific sub-regions. Finally, all the results and data available pre-dated COVID-19, and the impact of COVID-19 on testing rate or sexual behaviour is not included due to lack of data.

Our analysis suggests that the overall response to-date has contributed to the stabilisation of the syphilis outbreak in remote Indigenous communities of Australia. However, this response needs to be maintained for many years to come and additional responses that increase annual testing coverage are needed to reduce syphilis prevalence to pre-outbreak level within 5 years. Introducing community-wide testing on top of increases in annual testing coverage could hasten this reduction.

## Data Availability

The model is programmed using Java and is available at GitHub at https://doi.org/10.5281/zenodo.4057288.3

https://doi.org/10.5281/zenodo.4057288.

## Contributors

BBH implemented the model and performed simulations. BBH, RTG and DGR analysed the model output and prepared the first draft of the manuscript. RTG and DGR initiated the study and administered the project. RTG, RG and DGR acquired the project funding. DGR and ML provided the resources. BBH, DGR, RTG, RG and JSW conducted validation of data and results. All authors contributed to the conceptualisation of the study, reviewed drafts and approved the final manuscript.

## Declaration of interest

We declare no completing interests.

## Acknowledgements

The Kirby institute is funded by the Australian Government Department of Health and is affiliated with the Faculty of Medicine, UNSW, Sydney. The views expressed in this project will not necessarily represent the position of the Australian government.

This project was funded to provide guidance to the Multi-Jurisdictional Syphilis Outbreak Working Group (MJSO) and the Australian Health Protection Principle Committee (AHPPC) Enhanced Response Governance Group (the Governance Group) to assist in the roll-out of the national Enhanced Response and the development of activity work plans in Aboriginal Community Controlled Health Services and other settings. The MJSO and Governance Group provided surveillance data, guidance and feedback on the modelling methods and characteristics of the scenarios implemented.

The working group consists of the following members:

## Governance Group

- Prof Brendan Murphy, Chief Medical Officer (CMO) for the Australian Government;
- Ms Celia Street, First Assistant Secretary, Office of Health Protection (OHP), Department of Health (DoH);
- Dr Nathan Ryder, Chair of Multi-jurisdictional Syphilis Outbreak (MJSO) Working Group;
- Dr Dawn Casey, National Aboriginal Community Controlled Health Organisation (NACCHO);
- Dr Russell Waddell, South Australia Department of Health;
- Dr Sonja Bennett, Queensland Department of Health;
- Dr Hugh Heggie, Northern Territory Department of Health;
- Dr Paul Armstrong, Western Australia Department of Health;
- Prof James Ward, University of Queensland

## MJSO – represented organisations

- Australian Government Department of Health
- National Aboriginal Community Controlled Health Organisation
- Aboriginal Medical Services Alliance Northern Territory
- Northern Territory Department of Health
- Queensland Aboriginal and Islander Health Council
- Queensland Department of Health
- Cairns and Hinterland Health and Hospital Service (Qld)
- Townsville Health and Hospital Service (Qld)
- Aboriginal Health Council of Western Australia
- Western Australia Department of Health
- Pilbara Public Health Unit (WA)
- Kimberley Public Health Unit (WA)
- Aboriginal Health Council of South Australia
- South Australia Health and Medical Research Institute
- South Australia Department of Health
- Kirby Institute, UNSW Sydney
- Victorian Department of Health and Human Services
- NSW Ministry of Health

## Notes

### Competing Interest Statement

The authors have declared no competing interest.

### Author Declarations

As it is a modelling study using publicly available data no IRB approval is needed.

