## Appendix for "Evaluating strategies to combat a major syphilis outbreak in Australia among Aboriginal and Torres Strait Islander peoples in remote and regional Australia through mathematical modelling"

### Appendix: Syphilis Model

#### 1. Summary

An individual-based model of syphilis transmission is developed for this study. The model population of consists of 10550 heterosexual individuals spanning over 11 hypothetical locations. The first location constitutes the home for 5550 individuals and is denoted as the regional centre. The remaining 10 locations are home to 500 individuals each and denote the remote locations for the population.

The locations are linked as complete graph such that residents at each location are free to move away for a short period before returning home. See Section 2, "**Mobility**" for details on how mobility is implemented in the model.

The model population includes individuals of both genders of age 16-35, with the age, infection status, physical location, sexual partner history and sexual behaviour of individuals are tracked and updated on a daily basis.

The population distribution for each age and gender group is maintained throughout the simulation runs, with aged-out individuals replaced by a new individual of same gender and of age 15. The distribution is based on the estimated population size of Aboriginal and Torres Strait Islander Australians living in remote regions at 2010 by Australian Bureau of Statistics [1] and is shown in Table A1.

| Gender/age group | Percentage of population (%) |
|---|---|
| Male, 15-19 | 14.3 |
| Male, 20-24 | 13.6 |
| Male, 25-29 | 12.0 |
| Male, 30-34 | 10.0 |
| Female, 15-19 | 13.5 |
| Female, 20-24 | 13.7 |
| Female, 25-29 | 12.4 |
| Female, 30-34 | 10.4 |

*Table A1: The population distribution used in the model*

Individuals can form sexual partnership while they are at the same physical location. Partnership are maintained even if one or both partners have moved away, but sexual contact can only occur if both partners are at the same location at the same time. The frequency of partner change and duration of partnership is determined by the partner acquisition rate calculated based on the number of partners an individual can have in last 12 months. See Section 3, "**Sexual behaviour**" for the descriptions and parameters on how partnership are from are formed in this model.

Transmission of infection can occur if a sexual act occurs between an infectious individual and a susceptible individual. Infected individuals are considered as infectious if they are in the exposed, primary, secondary, early latent or recurrent stages of syphilis. If left untreated, individuals will eventually reach tertiary syphilis and remain there until they receive treatment. See Section 4, "**Infection (syphilis)**" for descriptions on how syphilis transmission defined in this model.

Individual can be tested through annual STI testing, and treatment is scheduled if infection is found. The model allowed some time delays between testing and treatment or could missed treatment entirely. See Section 5, "**Annual STI screening**" for descriptions and parameters governing the testing and treatments of individuals in this model.

The model is calibrated against the increase of infectious syphilis notification rate from 2013 to 2019 [2]. Each simulation run starts with burn-in period of 55 years (consists of 50 years with no infection, followed by the introduction of syphilis and then continues for another 10 years). 5000 simulations are then run and 100 simulations with the notification rate (define as the diagnostic rate from screening and testing from the model) match closest to the reported notification rate from 2013 to 2019 (as determined by smallest sum of squares difference) are selected as the baseline model for this study. Figure A1 shows a summary of the infectious syphilis notification rate from selected simulation runs against the notification rate from the surveillance report.

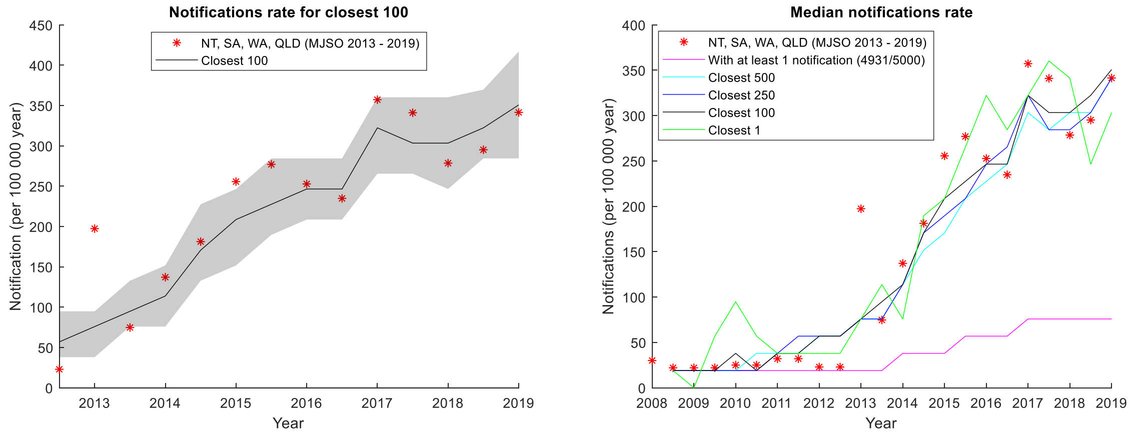


*Figure A1: Left: Infectious syphilis notification rate from 2010 – 2018 (red asterisk), against the infectious syphilis notification rate from 100 model simulations result (out of 5000) that have the closest match with notification rate from 2013 to 2019. The solid black line and the shaded region are the median and the $25^{th}$-$75^{th}$ percentile of those 100 simulation results respectively. Right: Infectious syphilis notification rate from 2010 – 2019 (red asterisk), against the median notification rate of infectious syphilis generated by the model by selecting: all simulation runs with at least 1 notification (pink), closest 500 (cyan), closest 250 (blue) and closest 100 (black).*

The model is programmed using Java and is available at GitHub at https://doi.org/10.5281/zenodo.4057288.[3]

#### 2. Mobility

The model includes the effects of on temporary mobility between multiple small populations. Mobility entails the movement of individuals away from their home location to a new location. Individuals stay at the new location for a predetermined period before returning to their home location. We assume individual stays away from home 2 weeks to 6 months, based on the finding from Prout [4].

The proportion of the population away from home is based on the findings reported in the study by Biddle and Prout [5] and is shown in Table A2.

| | Male | | | | Female | | | |
|---|---|---|---|---|---|---|---|---|
| Age | 16-19 | 20-24 | 25-29 | 30-35 | 16-19 | 20-24 | 25-29 | 30-35 |

| | | | | | | | | |
|---|---|---|---|---|---|---|---|---|
| Regional | 8% | 10% | 9% | 8% | 8% | 7% | 6% | 5% |
| Remote | 11% | 10% | 9% | 9% | 11% | 10% | 9% | 8% |

*Table A2: Percentage of individual away from home, in term of gender and home location.*

At each time step, the proportion of individuals away from home is calculated for each of the gender-age group defined in Table A2. If, for a particular gender-age group, the proportion away is less than the corresponding value listed in Table A2, then some individuals at home will be randomly selected and move away from home in the next time step, with their destination randomly selected from the 20 non-home locations. The number of individuals to move is calculate as the product of the differences between the proportion away and values listed in Table A2, and the total number of individuals within the particular gender-age group. This ensures the proportion of individuals away in the model to match closely with the data at every time step as Figure A2 shows. Note that there are more variations (i.e. more outliers, wider box etc) for the remote location due to smaller population size.

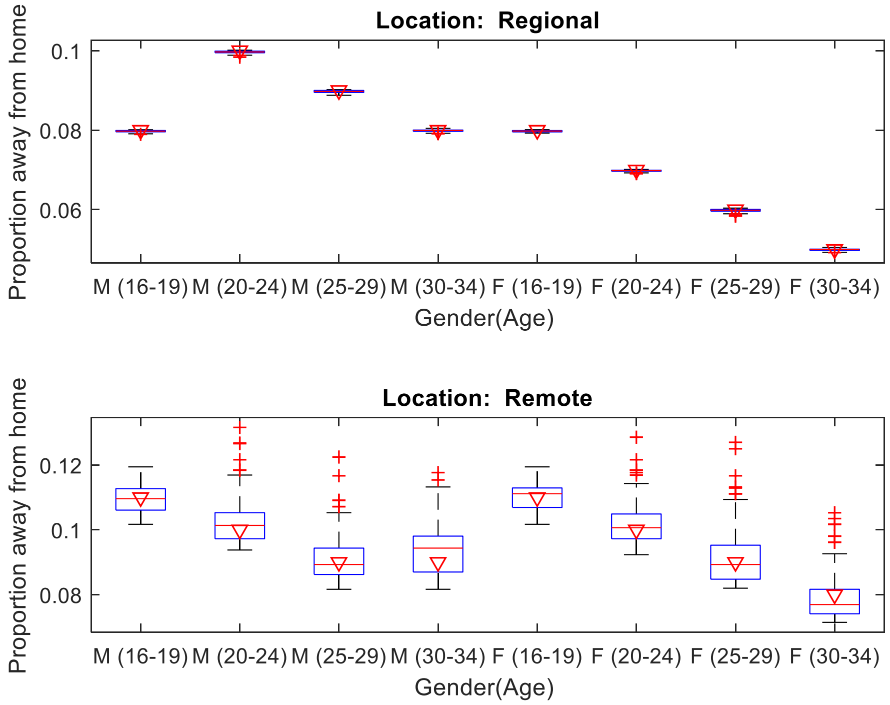


*Figure A2: Proportion of population away from home. The boxplot is the result from 5000 simulation runs of the baseline model. The triangles are the age, gender and location specific proportion of population away from home from Biddle and Prout (i.e. Table A2)*

##### 3. Sexual behaviour

The formation and dissolution of partnership is governed by the number of partners in last 12 months report from GOANNA [6] and is listed in Table A3.

| Number of partners in last 12 months | Age |
|---|---|

| | 16-19 | 20-24 | 25-29 | 30-35[1] |
|---|---|---|---|---|
| 0 | 9% | 7% | 9% | 9% |
| 1 | 40% | 47% | 55% | 62% |
| 2-4 | 42% | 38% | 32% | 27% |
| 5+ | 9% | 8% | 4% | 2% |

*Table A3: The relationship between age and the number of partners an individual has in last 12 months. The percentage represents the proportion of individuals within age group (i.e. columns) that have specified number of partners in last 12 months.*

The partnership status of all individuals in the modelled population is updated daily. At each one-day cycle, the number of partners in last 12 months for each individual is calculated. The number of individuals that seek or break partnership is then calculated by comparing the differences between value from the model and from the Table A3.

For example, let assume at a particular time step, the percentage of 16-19 years old that have 0, 1, 2-4, and 5+ partner in last 12 months are 8%, 43%, 40%, 9% respectively. It means there is an excess of the individuals of age 16-19 who has 1 partner in last 12 months. In order to ensure the model matches with data, some of them need to seek another partner (at probability of $\frac{(42-40)+(9-9)}{3} = \frac{2}{3}$) while the rest need to break their current partnership (at probability $\frac{9-8}{3} = \frac{1}{3}$). In this example, the number of individuals that seek or break partnership will be the 3% of the total number of individuals of age 16-19. That number of individuals is then randomly selected from those who has 1 partner in last 12 months, and the selected individual is to seek or break partnership according to probability calculated above. This ensures the partner seeking behaviour in the model to match closely with the data as Figure A3 demonstrates.

Sexual acts (required for transmission of infection) occur between two individuals within an existing partnership at the same physical location. In the model, the frequency of acts is 1.4 per week, based on the number of sexual acts per week reported in ASHR2[7]. The condom usage is 0.54 per acts as reported in GONNA [6].

[1] Since the upper age limit for GOANNA is 30, we estimate the number of partners from last 12 months for the 30-35 age group by extrapolating a linear curve across the 3-age group.

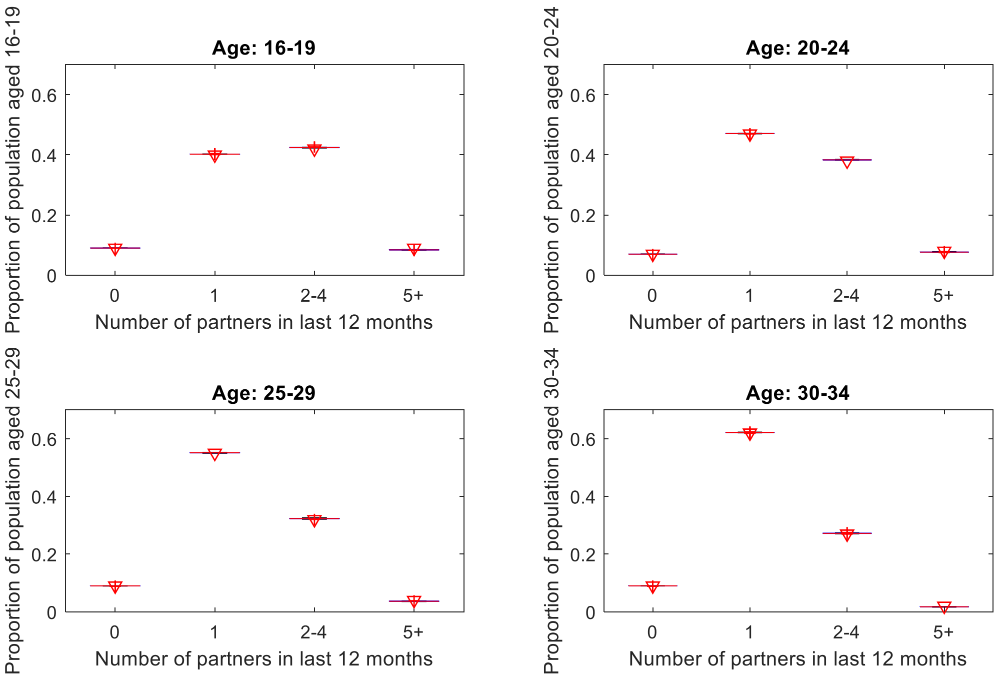


*Figure A3: The age specific distribution on the number of partners an individual has in last 12 months. The boxplot is the result from 100 simulation runs of the baseline model. The triangles are the data from GOANNA (see Table A3)*

*Sexual behaviour of travellers*

The model assumes some travellers with an existing partnership in their home location can have sexual contact with non-partners while away from home. While this is a fair assumption, reliable data on this factor is unavailable (and possibly never available). In this model it assumes that, on top of normal partner seeking pathway as described above, some traveller can seek one-off sexual contact (i.e. no partnership formed and only engage in sex act for one day) while away from home, but only with individuals at the same location who are also travellers or are also seeking partners as the traveller at the same time. Calibration result on same model on gonorrhoea and chlamydia suggests that if 53%, 18%, 5% and 2% of travellers of age 16-19, 20-24, 25-29 and 30-35 respectively can engage with sex outside partnership, then the prevalence of chlamydia and gonorrhoea is around the same level as those found in the STRIVE baseline prevalence survey [8]. These parameters are also applied to the current model.

#### 4. Infection (syphilis)

The natural history of syphilis is assumed to be same as a previous model on syphilis in MSM population [9]. Infected individuals are considered as infectious if they are in the exposed, primary, secondary, early latent or recurrent stages of syphilis. If left untreated, individuals will eventually reach tertiary syphilis and remain there until they receive treatment.

Figure A4 is the schematic diagram showing the stages and disease progression of syphilis as described in the model, and the infection parameters used in the model is listed in Table A4.

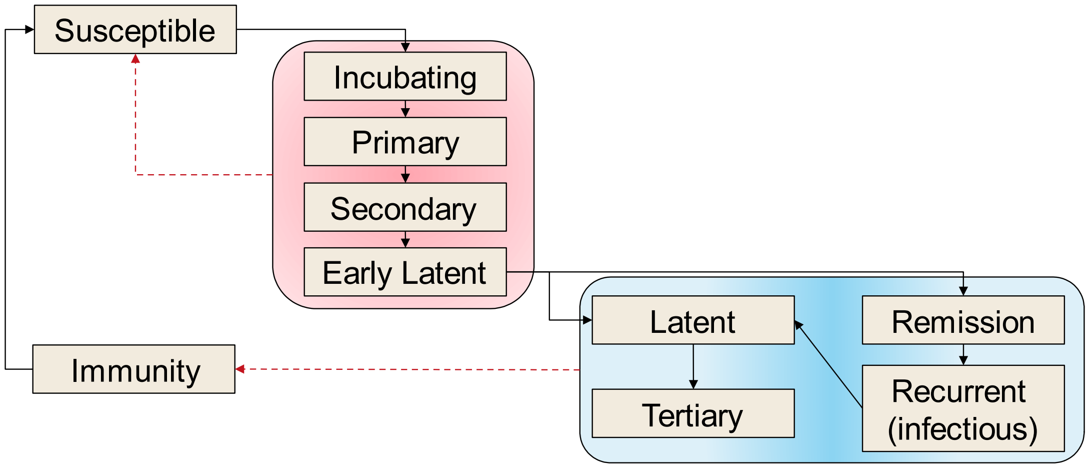


*Figure A4: Schematic diagram of stages and disease progression of syphilis in the model. (based on [9], figure 1).*

| **Parameters** | **Values** | **Source** |
|---|---|---|
| Duration of incubation period | 21 – 28 days | 9-12 |
| Duration of primary syphilis | 45 – 60 days | |
| Duration of secondary syphilis | 100 - 140 days | |
| Duration of early latent period[2] | 132 - 554 days | |
| Duration of latent period | 15 years | |
| Duration of remission | 6 months | |
| Duration of recurrent infection (infectious) | 90 days | |
| Duration of immunity from treatment | 5 years | |
| Transmission probability per act[3] | | |
| Male to female | 0.0525 | Calibrated |
| Female to male | 0.0430 | Assume to be 82% of transmission probability from male to female, based on the ratio from Johnson et al. [13] |
| Percentage of syphilis cases that recur after infectious syphilis | 25% | 9 |

*Table A4: Parameters related to syphilis infection*

When a sexual act occurs between an infectious individual and a susceptible individual, a uniform random number (range [0,1]) is generated, and transmission of infection to the susceptible individual will occur if that number is less than the transmission probability assigned for the simulation. The probability of developing symptoms is assigned by similar process. The duration of infection stages is sample from the uniform distributions with range as specified in Table A4, and is assigned on a per-infection basis.

[2] Total duration of infectiousness, which comprise of incubation, primary, secondary and early latent period, is believed to be 1-2 years

[3] It is assumed the transmission probability is halved if the infected

#### 5. Annual STI screening

*Pre-enhanced response (prior to mid-2013)*

*Annual testing coverage - remote*

For remote communities, it is assumed assume annual STI testing of individuals of age 16-35 are carried out in the population. The STI testing coverage varies according to gender and age and are based on baseline testing coverage from the STRIVE study and are listed in Table A5.

| Male (%) | | Female (%) | |
|---|---|---|---|
| 16-19 | 13 | 16-19 | 26 |
| 20-24 | 14 | 20-24 | 24 |
| 25-30 | 23 | 25-30 | 11 |
| 30-34 | 11 | 30-34 | 19 |

*Table A5: Annual STI screening coverage for remote locations prior to enhanced response at mid-2013*

*Annual testing coverage – regional*

Age and gender specific testing coverage data is not available from the GOANNA report or other sources. In this model, STI testing coverage in regional areas is assumed to have the same pattern as remote, but the values are adjusted by the overall testing coverage in GOANNA. In GOANNA, 44% surveyed in regional centre are tested in the last 12 months, compared to 48% surveyed in remote communities. Therefore, in this model we assumed the testing coverage for the regional centre is $\frac{0.44}{0.48} = 0.917$ of the coverage of residents in the remote communities. E.g. the testing coverage of 16-19-year-old males who reside in the regional centre will have a testing coverage of $0.13 \times \frac{0.44}{0.48} = 0.12$.

*Post-enhanced response (after mid-2013)*

Since the start of the outbreak we estimate that the overall level of syphilis testing in outbreak affected areas has increased substantially since 2013 from 30.1% per year in 2011 to 44.4% per year by the end of 2018 in 15-34-year-olds. This is based on the 6 months syphilis testing data from MJSO and is listed in Table A6.

As the coverage from MSJO included for remote region and only for age 15-24, the gender-age specific coverage used in the model set such that the coverage for all 15-24 in the remote communities in the model is the same as Table A6, while the coverage between each gender-age-location specific grouping has the same relative differences to the coverage prior to 2013. The testing cover use in the model is shown in Table A7

| **Year** | **Mid-2013** | **2014** | | **2015** | | **2016** | | **2017** | | **2018** | |
|---|---|---|---|---|---|---|---|---|---|---|---|
| **Testing coverage for age 15-24 (%)** | 35 | 34 | 36 | 40 | 41 | 47 | 46 | 48 | 49 | 52 | 52 |

*Table A6: Syphilis testing coverage for age 15-24 from mid-2013 to 2018, as supplied by MJSO.*

| **Year** | **Mid-2013** | **2014** | | **2015** | | **2016** | | **2017** | | **2018** | |
|---|---|---|---|---|---|---|---|---|---|---|---|
| **Annual testing coverage for remote communities (%)** | | | | | | | | | | | |
| **Male 16-19** | 24 | 23 | 25 | 27 | 28 | 32 | 31 | 33 | 33 | 35 | 35 |

| | | | | | | | | | | | |
|---|---|---|---|---|---|---|---|---|---|---|---|
| **Male 20-24** | 26 | 25 | 27 | 29 | 30 | 35 | 34 | 35 | 36 | 38 | 38 |
| **Male 25-30** | 20 | 20 | 21 | 23 | 24 | 27 | 26 | 28 | 28 | 30 | 30 |
| **Male 30-34** | 20 | 20 | 21 | 23 | 24 | 27 | 26 | 28 | 28 | 30 | 30 |
| **Female 16-19** | 48 | 46 | 49 | 55 | 56 | 64 | 62 | 66 | 66 | 71 | 70 |
| **Female 20-24** | 44 | 43 | 45 | 51 | 52 | 59 | 58 | 61 | 61 | 65 | 65 |
| **Female 25-30** | 42 | 41 | 44 | 48 | 50 | 57 | 55 | 58 | 59 | 63 | 62 |
| **Female 30-34** | 35 | 34 | 36 | 40 | 41 | 47 | 46 | 48 | 48 | 52 | 51 |
| **Annual testing coverage for regional centre (%)** | | | | | | | | | | | |
| **Male 16-19** | 22 | 21 | 23 | 25 | 26 | 29 | 29 | 30 | 30 | 32 | 32 |
| **Male 20-24** | 23 | 23 | 24 | 27 | 28 | 32 | 31 | 32 | 33 | 35 | 35 |
| **Male 25-30** | 18 | 18 | 19 | 21 | 22 | 25 | 24 | 25 | 26 | 27 | 27 |
| **Male 30-34** | 18 | 18 | 19 | 21 | 22 | 25 | 24 | 25 | 26 | 27 | 27 |
| **Female 16-19** | 44 | 42 | 45 | 50 | 51 | 59 | 57 | 60 | 61 | 65 | 64 |
| **Female 20-24** | 40 | 39 | 42 | 46 | 47 | 54 | 53 | 55 | 56 | 60 | 59 |
| **Female 25-30** | 39 | 37 | 40 | 44 | 46 | 52 | 51 | 53 | 54 | 57 | 57 |
| **Female 30-34** | 32 | 31 | 33 | 37 | 38 | 43 | 42 | 44 | 44 | 47 | 47 |

*Table A7: Age-gender syphilis testing coverage used in the model from mid-2013 to 2018 (prior to adjustment to testing coverage)*

*Treatment sensitivity and delay*

Table A8 listed other input parameters related to the sexual behaviour in the modelled population.

| **Parameters** | **Value** | **Source** |
|---|---|---|
| Test sensitivity | 0.98 | Assumption |
| Treatment delay[4] (Remote) | | |
| None | 27.4% | TTANGO[5] |
| 2 days or less | 30.1% | |
| 7 days or less | 47.2% | |
| 4 months or less | 85.7% | |
| Treatment delay (Regional) | 90% treated within 7 days | Assumption |

*Table A8: Parameters related to testing*

It is assumed that treatment will be dictated by the location where the test is carried out. For example, if an individual tested positive at home location and is scheduled to be treated 7 days later but was not at home at that time, then treatment will not occur for that individual.

#### 6. Adjustment to baseline testing coverage

As described in the main text, we assumed reported testing coverage, calculated from raw testing data as described in Table A7 above, is not the exact representation of testing coverage in the population. We have

[4] It is assumed the patient will not be treated if that patient has not received treatment after 4 months.
[5] Personal communication with Louise Causer (investigator on the TTANGO study).

identified that there are potential under counting in number of tests from private laboratories, and over counting due to tests in visitors from outside the outbreak affected areas and multiple tests in individuals. After consultations with experts in the field, we estimated a more accurate testing coverage should be 75% of the testing coverage estimated based on number of tests carried out. Hence for the results shown in the main manuscript, a 75% adjustment was in place for the testing coverage recorded during enhance response (i.e. between mid-2013 to 2018).

Figure A5 and Figure A6 below show the outcomes under each of the scenario described in the manuscript should the adjustment to testing coverage is not made. They suggested that, should the adjustment to testing coverage has not been made, infectious syphilis prevalence is already on the decline without additional increase in testing coverage.

Note that the time of writing (early 2020), while the number syphilis notifications over the entire affected region remain stable, this is not uniform across different jurisdictions and regions, with some regions recorded increase in notifications while other recorded decreases. Given an overestimation of testing coverage from raw testing data is more likely than an underestimate, we have decided to take a conservative approach, and used the adjusted testing coverage as the main result for the manuscript.

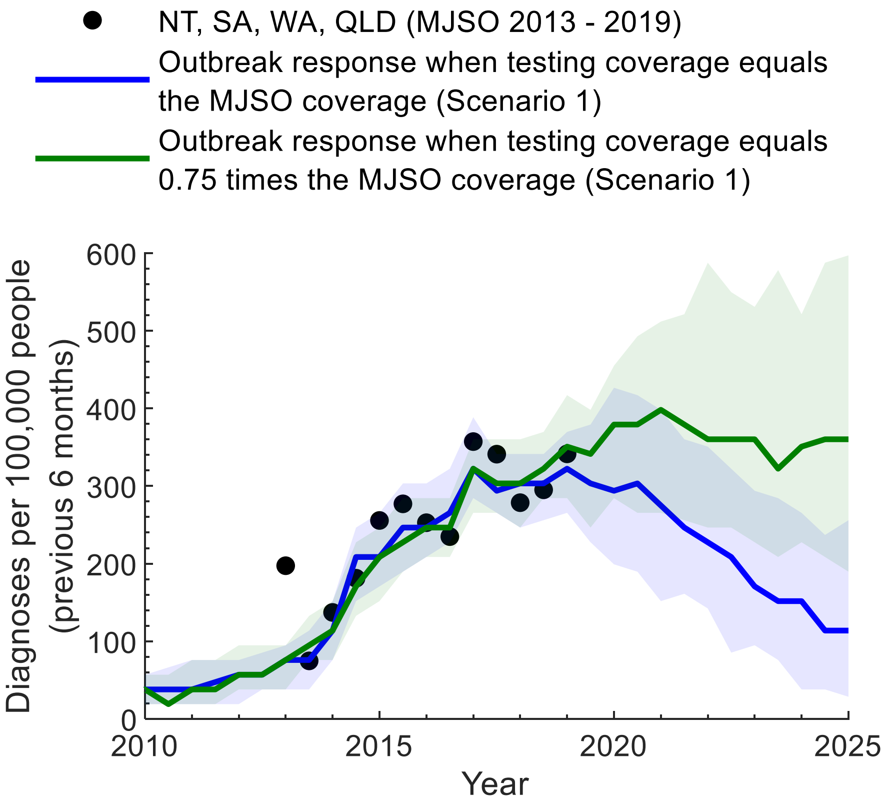


*Figure A5: Estimated diagnoses from the model for the two main testing coverage assumptions compared to notifications data. Diagnoses per 100,000 people from 2010 to 2025 under the current response (status-quo) scenario compared to the total notification rate in the outbreak affected areas (black points). The solid line and the shaded region are the median and interquartile range, respectively, from the 100 selected model runs that best fit the data.*

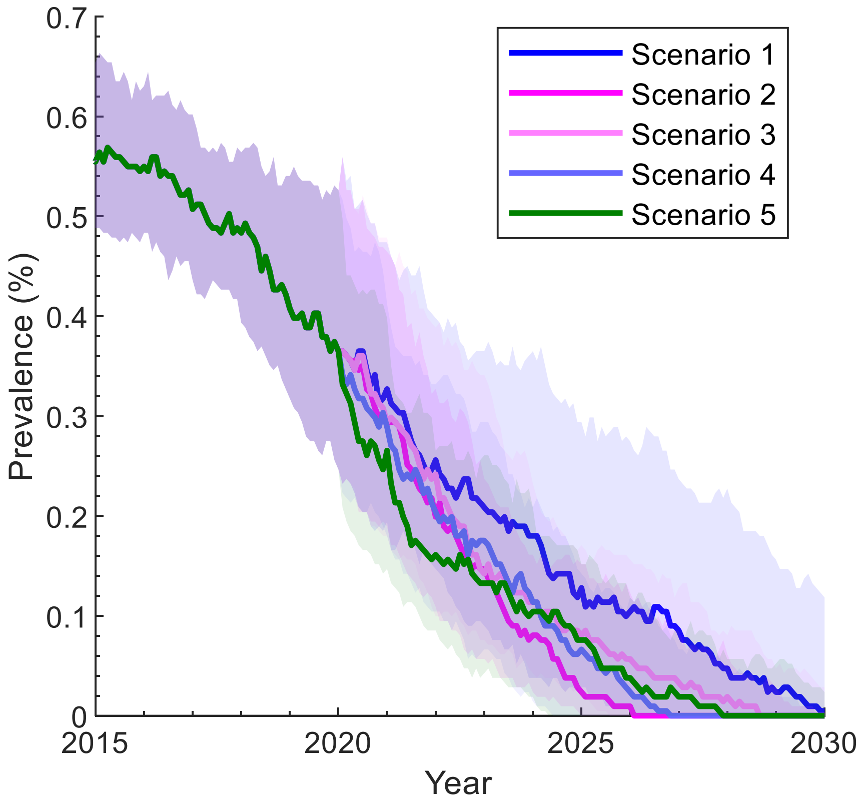


*Figure A6: Infectious syphilis prevalence trajectories for Scenarios 1 -5 over the period 2015-2030 under the assumption that testing coverage equals MJSO testing coverage. The solid lines and shading are the median and interquartile range, respectively, from 100 model runs that provide the closest match (in terms of smallest summed square differences) with the notification rate from 2013 to the end of 2019.*

#### 7. Effect of earlier response

In the manuscript, we assume further expansion of testing interventions can only occur at the end of 2019, when infectious syphilis has already spread to multiple judications and outbreak is already peaked in some communities. Given the first sign of a possible syphilis outbreak is detected in the communities back in early 2010's, there is interest in examining what could have happened if expansion of testing intervention can happen before a full outbreak has taken place.

In this section, we examine the possible benefit of increasing annual testing coverage to 60% within 2 years (i.e. Scenario 2 in the manuscript), but at end of 2013 and 2011 instead of 2019. Figure A7 compares the Infectious syphilis prevalence trajectories under each scenario. The results suggest that while increasing testing coverage earlier alone cannot stop the rise of infectious syphilis prevalence immediately, it does lead to a smaller peak prevalence (<0.7% if introduced at the end of 2013, <0.4% if introduced at the end of 2011), and would have return to pre-outbreak level (of less than 0.2%) by 2017. Introducing the increase in testing coverage earlier instead of end of 2019 has led to more than 500 infections averted, as the results in Table A9 shows.

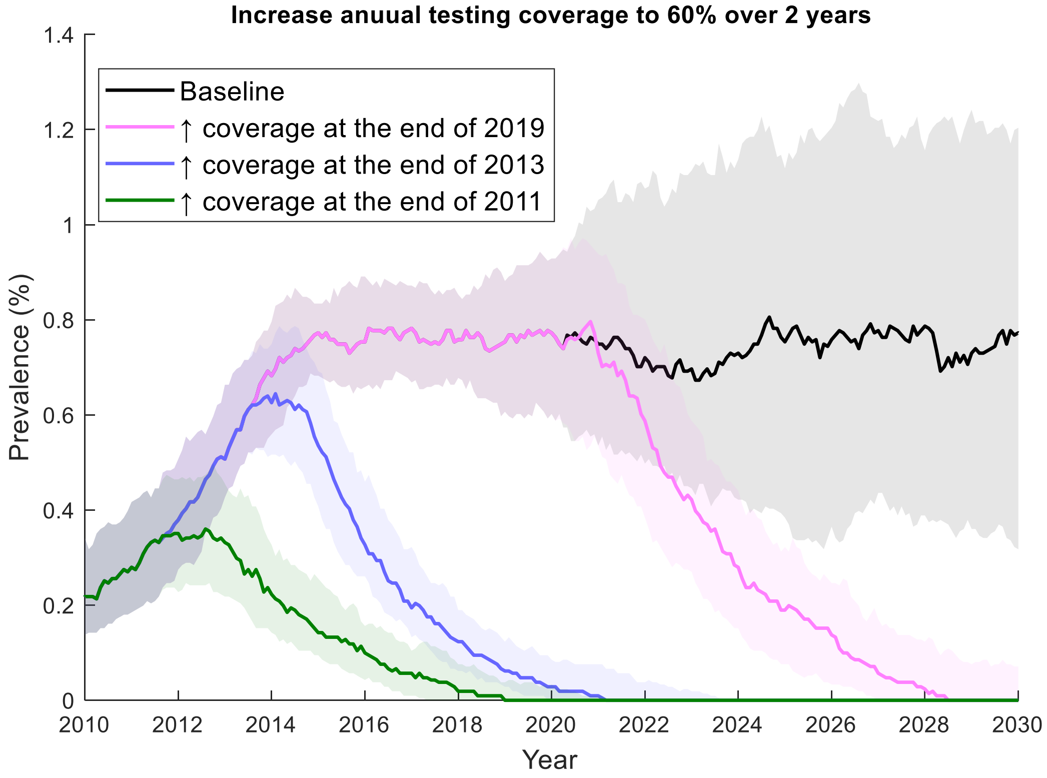


*Figure A7: Infectious syphilis prevalence trajectories over the period 2015-2030 under baseline model (Scenario 1 in main text, black), increase annual testing coverage to 60% over 2 years at the end of 2019 (Scenario 2 in the main text, magenta), and the increase is brough forward to end of 2013 (blue) and end of 2011 (green). The solid lines and shading are the median and interquartile range, respectively, from 100 model runs that provide the closest match (in terms of smallest summed square differences) with the notification rate from 2013 to the end of 2019.*

| **Scenario** | **Number of new case (2011-2044)** | **Infection averted (2011-2044)** | | **Cumulative notification (2011-2044)** |
|---|---|---|---|---|
| | | **from baseline** | **from Scenario 2** | |
| Counterfactual scenario where testing coverage remains at 2013 levels. (Scenario 0) | 7333.0 (6709.0 - 7858.5) | < 0 | < 0 | 1711.0 (1559.5 - 1872.0) |
| Baseline - Annual testing coverage remains at the levels at the end of 2019 (Scenario 1) | 2497.5 (1360.0 - 3558.5) | Ref. | < 0 | 1165.5 (613.0 - 1704.5) |
| Annual testing coverage is increased to 60% over a period of 2 years from the end of 2019 (Scenario 2) | 935.0 (757.0 - 1150.0) | 1480.0 (368.0 - 2410.0) | Ref. | 489.5 (380.5 - 648.5) |
| Annual testing coverage is increased to 60% over a period of 5 years from the end of 2019 (Scenario 3) | 1119.0 (854.0 - 1331.5) | 1366.0 (311.0 - 2080.5) | < 0 | 582.0 (429.5 - 774.0) |
| Annual testing coverage remains at 2019 level + 2 community screening of 30% (Scenario 4) | 2300.5 (1347.0 - 3187.5) | 16.5 (-635.0 - 854.0) | < 0 | 1117.5 (633.0 - 1551.5) |
| Annual testing coverage is increased to 60% over a period of 2 years from end of 2019 + 2 community screening of 30% (Scenario 5) | 892.0 (724.5 - 1034.0) | 1643.5 (446.5 - 2501.5) | 53.5 (-45.0 - 173.5) | 463.5 (345.0 - 575.5) |
| Annual testing coverage is increased to 60% over a period of 2 years from the end of 2013 | 373.5 (275.0 - 452.0) | 2118.5 (963.5 - 3141.5) | 590.5 (406.5 - 778.5) | 220.5 (170.0 - 284.5) |
| Annual testing coverage is increased to 60% over a period of 2 years from the end of 2011 | 189.5 (114.5 - 285.0) | 2272.5 (1121.0 - 3350.5) | 702.0 (582.0 - 950.5) | 143.0 (93.5 - 198.5) |
| Annual testing coverage is increased to 60% over a period of 2 years from the end of 2011 + 2 community screening of 30% coverage | 166.0 (95.5 - 250.0) | 2195.5 (1162.5 - 3368.0) | 732.0 (591.0 - 991.0) | 114.5 (75.0 - 177.0) |
| Annual testing coverage is increased to 60% over a period of 2 years from the end of 2011+ 5 community screening of 30% coverage | 168.5 (87.0 - 238.5) | 2280.5 (1193.5 - 3340.5) | 733.0 (595.0 - 964.5) | 122.5 (80.0 - 176.5) |

| | | | | |
|---|---|---|---|---|
| Annual testing coverage is increased to 60% over a period of 2 years from the end of 2011 + 2 community screening of 60% coverage | 156.5 (79.0 - 222.0) | 2307.5 (1178.5 - 3317.5) | 767.5 (616.5 - 997.5) | 121.0 (67.5 - 170.5) |
| Annual testing coverage is increased to 60% over a period of 2 years from the end of 2011 + 5 community screening of 60% coverage | 145.0 (65.5 - 201.5) | 2286.0 (1161.5 - 3369.0) | 761.5 (633.5 - 1000.0) | 111.5 (65.0 - 165.0) |

*Table A9: The number of new syphilis cases and notifications across the 2011-2044 period as predicted by the model under scenarios listed in the manuscript, in comparison to scenarios where the increase of annual screening coverage is introduced during earlier stage of syphilis outbreak.*
